# Revealing the influence of national public health policies for the outbreak of the SARS-CoV-2 epidemic in Wuhan, China through status dynamic modeling

**DOI:** 10.1101/2020.03.10.20032995

**Authors:** Tianyi Qiu, Han Xiao

## Abstract

**Background:** The epidemic caused by SARS-CoV-2 was first reported in Wuhan, China, and now is spreading worldwide. The Chinese government responded to this epidemic with multiple public health policies including locking down the city of Wuhan, establishing multiple temporary hospitals, and prohibiting public gathering events. Here, we constructed a new real-time status dynamic model of SEIO (MH) to reveal the influence of national public health policies and to model the epidemic in Wuhan.

**Methods:** A real-time status dynamic model was proposed to model the population of Wuhan in status Susceptible (S), Exposed (E), Infected with symptoms (I), with Medical care (M), and Out of the system (O) daily. Model parameters were fitted according to the daily report of new infections from Jan. 27^th^, 2020 to Feb. 2^nd^, 2020. Using the fitted parameters, the epidemic under different conditions was simulated and compared with the current situation.

**Finding:** According to our study, the first patient is most likely appeared on Nov. 29^th^, 2019. There had already been 4,153 infected people and 6,536 exposed ones with the basic reproduction number *R*_0_ of 2.65 before lockdown, whereas *R*_0_ dropped to 1.98 for the first 30 days after the lockdown. The peak point is Feb. 17^th^, 2020 with 24,115 infected people and the end point is Jun. 17^th^, 2020. In total, 77,453 people will be infected. If lockdown imposed 7 days earlier, the total number of infected people would be 21,508, while delaying the lockdown by 1-6 days would expand the infection scale 1.23 to 4.94 times. A delay for 7 days would make the epidemic finally out of control. Doubling the number of beds in hospitals would decrease the total infections by 28%, and further investment in bed numbers would yield a diminishing return. Last, public gathering events that increased the transmission parameter by 5% in one single day would increase 4,243 infected people eventually.

**Interpretation:** Our model forecasted that the peak time in Wuhan was Feb. 17^th^, 2020 and the epidemic in Wuhan is now under control. The outbreak of SARS-CoV-2 is currently a global public health threat for all nations. Multiple countries including South Korea, Japan, Iran, Italy, and the United States are suffering from SARS-CoV-2. Our study, which simulated the epidemic in Wuhan, the first city in the world fighting against SARS-CoV-2, may provide useful guidance for other countries in dealing with similar situations.

**Funding:** National Natural Science Foundation of China (31900483) and Shanghai Sailing program (19YF1441100).

**Research in context:** *Evidence before this study:* The epidemic of SARS-CoV-2 has been currently believed to started from Wuhan, China. The Chinese government started to report the data including infected, cured and dead since Jan 20^th^, 2020. We searched PubMed and preprint archives for articles published up to Feb 28^th^, 2020, which contained information about the Wuhan outbreak using the terms of “SARS-CoV-2”, “2019-nCoV”, “COVID-19”, “public health policies”, “coronavirus”, “CoV”, “Wuhan”, “transmission model”, etc. And a number of articles were found to forecast the early dynamics of the SARS-CoV-2 epidemic and clinical characteristics of COVID-19. Several of them mentioned the influence of city lockdown, whereas lacked research focused on revealing the impact of public health policies for the outbreak of SARS-CoV-2 through modeling study.

*Added value of this study:* As the first study systemically analysis the effect of three major public health policies including 1) lockdown of Wuhan City, 2) construction of temporary hospitals and 3) reduction of crowed gathering events in Wuhan city. The results demonstrated the epidemic in Wuhan from the potential first patient to the end point as well as the influence of public health policies are expected to provide useful guidance for other countries in fighting against the epidemic of SRAS-CoV-2.

*Implications of all the available evidence:* Available evidence illustrated the human-to-human transmission of SARS-CoV-2, in which the migration of people in China during the epidemic may quickly spread the epidemic to the rest of the nation. These findings also suggested that the lockdown of Wuhan city may slow down the spread of the epidemic in the rest of China.

## Introduction

In December 2019, a pneumonia case caused by the new coronavirus SARS-CoV-2 was firstly reported in Wuhan, the capital city of Hubei Province, China^1^. The serious clinical symptoms of the viral infection including fever, dry cough, dyspnea, and pneumonia, may result in progressive respiratory failure and even death^2^. The disease which quickly spread to the rest of China and other continents, was currently been named as COVID-19 by the World Health Organization (WHO) ^3,4^. Till March 1^st^, 2020, at least 79,971 confirmed cases and 2,873 death caused by SARS-CoV-2 were reported by the National Health Commission of China^5^.

The epidemiological study of SARS-CoV-2 infected pneumonia provided evidence of human-to-human transmission^6,7^. Subsequently, the Chinese government quickly responded to this epidemic with a series of public health policies. For example, Wuhan, the largest city in central China with over 9 million permanent residents, was blocked for both outside and internal transportation to contain the spread of the epidemic on Jan 23^rd^, 2020 ^8^. Meanwhile, 31 provinces, cities and autonomous regions launched the level 1 public health response before Jan 30^rd^, 2020^9^. Moreover, multiple converted hospitals including Huoshenshan hospital, Leishenshan hospital, and mobile cabin hospital were constructed or renovated to support the battle against the epidemic in Hubei, China^10^.

Currently, several epidemic models were established for SARS-CoV-2 and the basic reproductive number R0 was estimated from 2 to 5^11-14^. The rapid transmission at the early stages might have been accelerated because of the out-break time is close to the Chinese Lunar New Year. The positive news is, besides Hubei province, the daily increase of new laboratory-confirmed patients in other regions of China has been declining since the peak on Feb 3^rd^, 2020, 11 days after the lockdown of Wuhan and 4 days after level 1 public health response in other regions ^15^. This further indicated that timely public health policies might significantly prevent the rapid spread of the epidemic.

Here, we proposed a status dynamic Susceptible-Exposed-Infectious-Out of the system (with Medical care/maximum beds in Hospital) SEIO (MH) model, to reveal the influence of the national public health response for the SARS-CoV-2 epidemics outbreak in Wuhan, China. Different from the traditional Susceptible-Exposed-Infectious-Recovered (SEIR) continuous model^13^. SEIO(MH) model is a real-time discrete model with two more essential parameters: 1) the number of people with medical care (M), and 2) the currently available beds in hospitals. The local status dynamic SEIO (MH) model could simulate the population in each status at any date during the epidemic. Also, this model could simulate multiple national public health responses including: 1) lockdown of the traffic in one city, 2) the hospital bed shortage as well as the amelioration in the medical system when multiple converted hospitals are available, and 3) the temporary public gathering events.

According to the simulation results, the first patient (patient zero) was more likely to appear on Nov. 29^th^, 2019. From Nov. 29^th^, 2019 to Jan. 23^th^, 2020, the basic reproduction number *R*_0_ was estimated at 2.65 before the lockdown of Wuhan. Till Jan 22^th^, 2020 before lockdown, the cumulative number of infections and exposures was 4,153 and 6,536, respectively. After the lockdown, the value of *R*_0_ decreased to 1.98 for the first 30 days. The peak point was simulated on Feb. 17^th^, 2020 with 24,115 infected people and the endpoint would be near June 17^th^, 2020. The total number of infected people in Wuhan was predicted to be 77,453. Moreover, we also simulated the infections under the three aforementioned public health responses. The construction of this model could not only study the SARS-CoV-2 epidemic outbreak in Wuhan, but also help design appropriate national public health policies for other countries.

## Materials and Methods

### Dataset

The increasing number of infected, cured and dead from Jan 27^th^, 2020 to Feb 2^nd^, 2020 were derived from the Health Commission of Hubei Province ^16^. Migration index before the lockdown and after the lockdown was derived from BAIDU migration ^17^. Beds in hospitals were derived from the news and reports from the Wuhan government ^18^.

### Notation of Parameters

The following variables denote the total number of people in each of the following statuses:

- *S*: Susceptible
- *E*: Exposed (infected but without symptoms yet)
- *I*: Infected and with symptoms
- *M*: with Medical care
- *M*: maximum number of beds in Hospital
- *O*: Out of the system (cured/dead)

Let *T* be the time point, *S*(*T*) denotes the total number of susceptible people at the time *T*. As well as other variables including *E*(*T*), *I*(*T*), *M*(*T*), (*T*), and *O*(*T*). Δ denote the increase between two consecutive days. For example, Δ*S*(*T*) = *S*(*T*) − *S*(*T* − 1). As well as other variables including Δ*E*(*T*), Δ*I*(*T*), Δ*M*(*T*), Δ (*T*), and Δ*O*(*T*). Let a and f3 be the infection coefficients that *E* imposes on *S* and *I* imposes on *S*, respectively.

Also, we defined status-dependent parameter *Pr* to describe the status transmission probability. For example, *Pr*_*E*→*I*_(t) denotes the probability of a person transformed from status *E* to status *I* after t days. As well as other parameters including *Pr*_*M*→*O*_, *Pr*_*I*→*M*_ and *Pr*_*I*→*O*_.

Current infectious will be *I* + *M*, total infectious will be *I* + *M* + *O*, the peak time is defined as the date with the maximum of *I* + *M*, the turning point is defined as the date when *O* > *I* + *M*, the end point is defined as the date when *O* > 0.99 (*E* + *I* + *M* + *O*).

### Status dynamic model and parameter estimation

We denote *C* as the size of the population in a local area being considered and *γ* as the size of initial infections (a small integer). Given any status such as *E*(*T*) at the time T, Δ^+^ *E*(*T*) was the number of people entered into status *E* at the time *T*. At *T* = 0, *S*(0) = *C* and *E*(0) = *γ*. Also, (*T*) equals to the maximum number of beds in hospitals at the time *T*. The transition rules of four status including *E, I, M*, and *O* were illustrated in Figure 1. Definition of each status can be found in *Supplementary Methods*.

**Figure 1:**
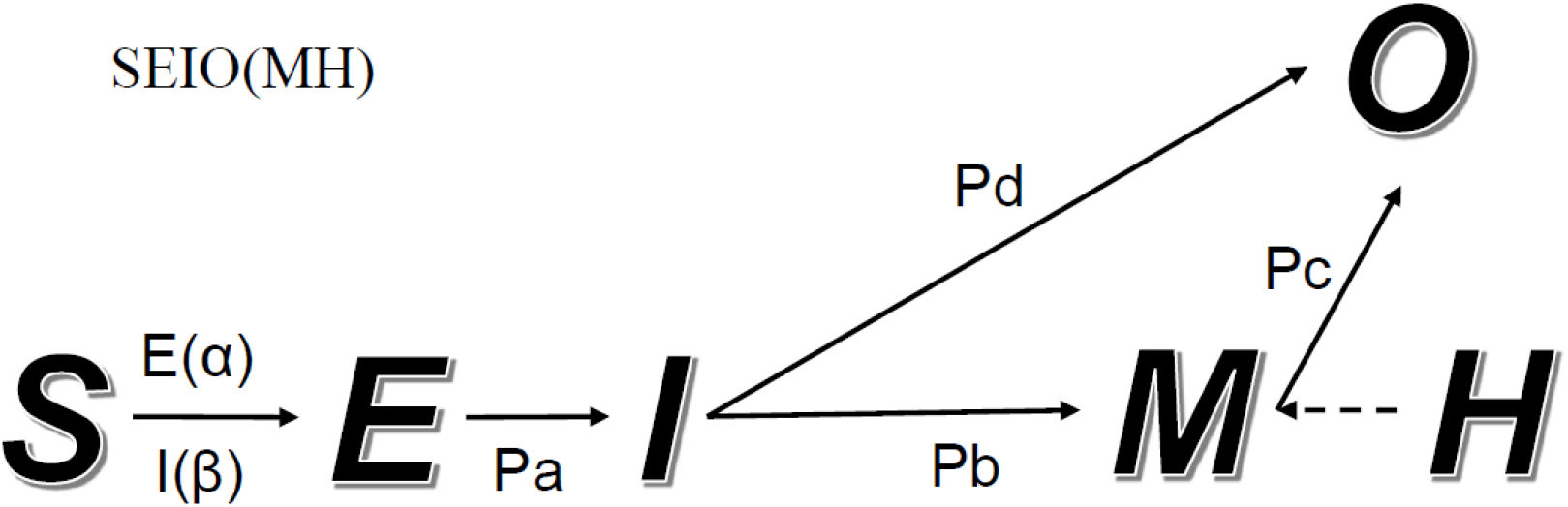
Illustration of the status dynamic SEIO (MH) model. S, E, I, M and O represent the people in the status of susceptible, exposed, infected with symptoms, with medical care and out of the system. The number of M is limited by the current number of H, which is the beds in hospitals. The arrow indicates the direction of status transmission. α and β represent the infection coefficients that E imposes on S and I impose on S, respectively. *P*_*a*_ to *P*_*d*_ represents the transmission probability between two statuses.

In SEIO (MH) modeling, status parameters of *S*(*T*_0_) *E*(*T*_0_), *I*(*T*_0_), *M*(*T*_0_), and *O*(*T*_0_), as well as transmission parameters of *α, β, Pr*_*E*→*I*_, *Pr*_*I*→*M*_, *Pr*_*I*→*O*_, and *Pr*_*M*→*O*_, were fitted based on the increasing number of people in each status reported by Health Commission of Hubei Province ^16^ during Jan. 27^th^, 2020 to Feb. 2^nd^, 2020.

## Results

### Model construction and status dynamic modeling of epidemic in Wuhan

The parameters of the status dynamic SEIO (MH) model were fitted according to the daily reports of new infections in Wuhan from Jan 27^th^ to Feb 2^th^, during which the daily new increase of people in status *I* was monotonously increasing with time. The initial number of people in status *I* and *E* were simulated from 500 to 10,000, while parameter *α* and *β* were simulated from 10^−9^ to 10^−11^, respectively. Also, the median time of people from status *M* to status *O* and people from status *I* to status *O* were simulated according to the increasing number of cured and dead from Jan 27^th^ to Feb 2^th^. By minimizing the absolute error to the ground truth number in status *I*, the statuses on Jan 27^th^ can be inferred.

Based on the fitted parameters, the simulation showed that it would take 14 days for people to transfer from status *M* to status *O*. For those out of the system before been hospitalized, it would take 28 days to transfer from status *I* to status *O*. Results also illustrated that at the initial date (*T*0) for fitting on Jan 26^th^, there were 3,998 symptomatic infections, 7,984 contacts, 3,815 hospitalizations and 696 out of the system (Cured or Dead) in Wuhan, in which the accumulative number of infected people (I+M+ O) raised up to 8,509 on Jan 26^th^.

With the default parameters, the epidemic in Wuhan was described by local status dynamic SEIO (MH) modeling. Illustrated in Figure 2a, patient zero was inferred to appear on Nov. 29^th^, 2019. From that day to Jan 23^th^, 2020, when less intervention was applied to prevent the epidemic, the number of infected people increased exponentially. After Jan 23^th^, the circulation intensity of people in Wuhan decreased sharply after the city lockdown ^19^. However, the people infected with symptoms, exposed and in hospitalization were estimated to be 1,682, 6,536 and 2,471 respectively on Jan 22^nd^, 2020 and increased to 1,999, 7,676 and 2,877 respectively on Jan 23^rd^, 2020 (Figure 2b), which was a large population with infected people and would lead to the outbreaks of the epidemic in the early period even after the lockdown.

**Figure 2:**
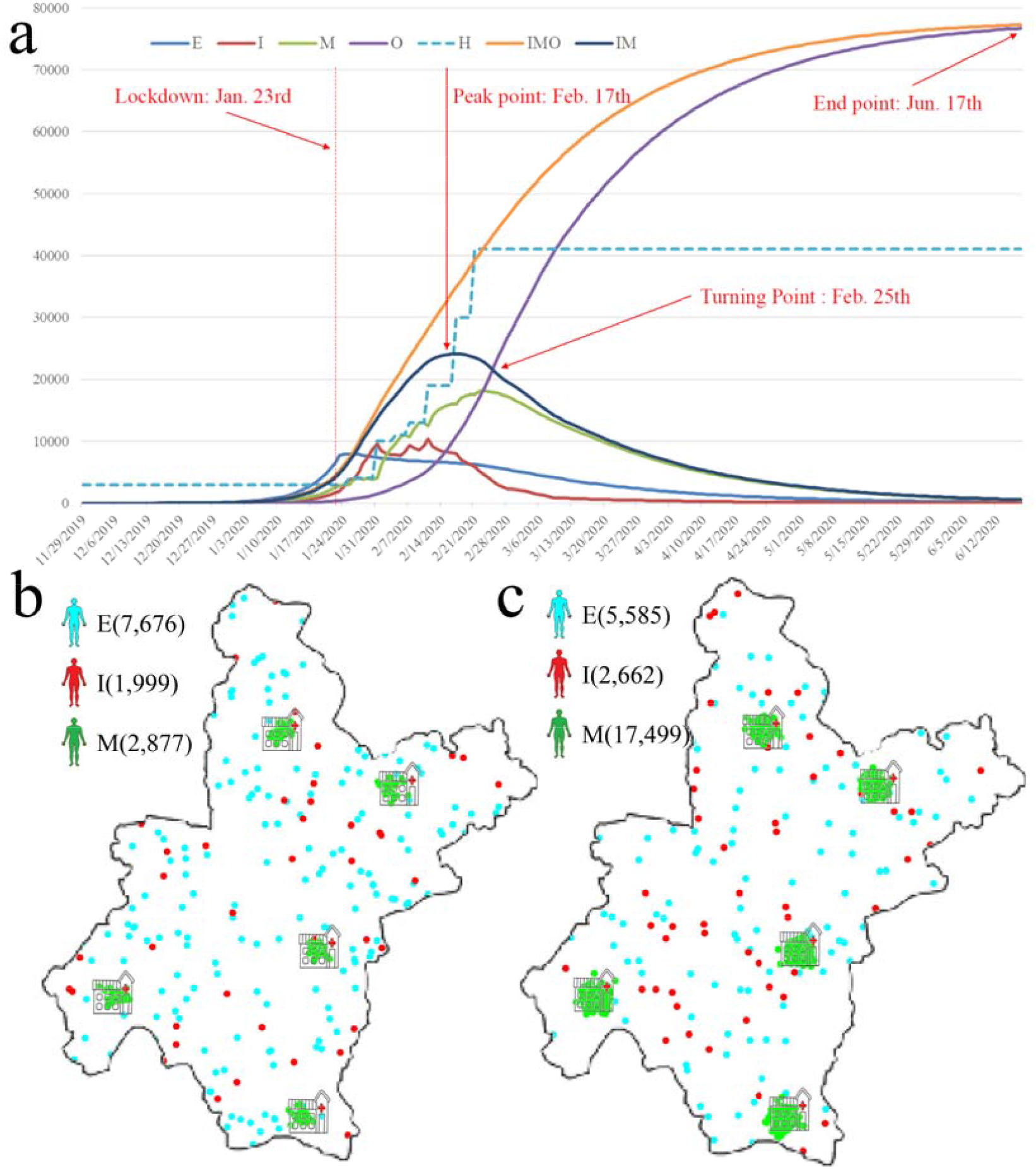
Simulation of the epidemic of SARS-CoV-2 in Wuhan. a) The epidemic of SARS-CoV-2 in Wuhan from patient 0 to end point. b) The epidemic situation at the time point of Wuhan lockdown, blue, red and green dots located on the map of Wuhan represent the exposed, infected with symptoms and with medical care respectively. c) The epidemic situation at peak time, blue, red and green dots located on the map of Wuhan represent the exposed, infected with symptoms and with medical care respectively.

Notably, the hospital was full of people (>80% of the current) in status *I* from Jan 22^th^, 2020 to Feb 10^th^, 2020. After the increasing number of beds in hospitals at Date Jan 25^th^, Jan 31^th^, Feb 6^th^, and Feb 10^th^, the hospital was still full immediately, indicating the deficiency of medical resources in Wuhan at the early period (Figure 2a). After the large scale of mobile cabin hospitals, the shortage of beds in hospitals was improved. We expected that the peak point in Wuhan would be Feb 17^th^, 2020 with 24,115 infected people, including 7,676 exposed, 1,999 infected with symptoms and 2,877 with medical care (Figure 2c), and the epidemic would end on Jun. 17^th^, 2020 with 77,453 infected people eventually, which was basically consistent with Yang’s study ^14^. The basic reproduction number was estimated to be 2.65 and would drop to 1.98 after one month since the lockdown. This result suggested that the choice of lockdown date and the number of hospital beds would significantly affect the development of the epidemic. Also, it should be noticed that the time of the epidemic period is overlapped with the Chinese Lunar New Year, in which public gathering events are expected. Thus, 1) the date of lockdown, 2) the number of available hospital beds 3) the public gathering events, were considered in our simulation.

### Impact of different public health policies

#### Impact of the lockdown of Wuhan

Here, we studied the effect of advancing or delaying the lockdown date by 1 to 7 days. Results indicated that, if Wuhan was blocked on Jan 16^th^, the accumulated infectious would be 4,063 on Jan 22^th^, which was slightly below 4,521 on the same date in current status. However, the exposed people on Jan 22^th^ were 2,427, only 37% of the simulated number (2,428/6,536) with the current lockdown date. Moreover, the peak time and endpoint would be advanced by 10 days (Figure 3). The total number of infected people would be 21,508, 28% (21,508/77,453) of the current situation and even one-day advance would cause only 82% (63,226/77,453) infections at the end point (Table 1).

**Table 1.** Simulation results of Wuhan city lockdown with 1-7 days advance or delay

| Lockdown<sup>a</sup> | R0 before<sup>b</sup> | R0 after<sup>c</sup> | End point<sup>d</sup> | Infected (E)<sup>e</sup> | Peak time<sup>f</sup> | Turning time<sup>h</sup> |
| --- | --- | --- | --- | --- | --- | --- |
| 2020-01-16 | 2.648 | 1.947 | 09/06/20 | 21,508 | 07/02/20 | 15/02/20 |
| 2020-01-17 | 2.650 | 1.953 | 10/06/20 | 25,868 | 11/02/20 | 17/02/20 |
| 2020-01-18 | 2.648 | 1.960 | 12/06/20 | 31,743 | 12/02/20 | 19/02/20 |
| 2020-01-19 | 2.649 | 1.965 | 13/06/20 | 38,199 | 13/02/20 | 20/02/20 |
| 2020-01-20 | 2.649 | 1.968 | 14/06/20 | 45,467 | 14/02/20 | 21/02/20 |
| 2020-01-21 | 2.650 | 1.970 | 14/06/20 | 53,654 | 14/02/20 | 22/02/20 |
| 2020-01-22 | 2.650 | 1.971 | 15/06/20 | 63,226 | 15/02/20 | 23/02/20 |
| 2020-01-23 | 2.651 | 1.980 | 17/06/20 | 77,453 | 17/02/20 | 25/02/20 |
| 2020-01-24 | 2.652 | 1.988 | 18/06/20 | 95,273 | 20/02/20 | 27/02/20 |
| 2020-01-25 | 2.652 | 1.992 | 19/06/20 | 114,105 | 22/02/20 | 29/02/20 |
| 2020-01-26 | 2.652 | 2.003 | 20/06/20 | 141,409 | 23/02/20 | 02/03/20 |
| 2020-01-27 | 2.657 | 2.014 | 22/06/20 | 176,300 | 25/02/20 | 04/03/20 |
| 2020-01-28 | 2.659 | 2.027 | 24/06/20 | 222,384 | 27/02/20 | 06/03/20 |
| 2020-01-29 | 2.659 | 2.081 | 20/07/20 | 382,695 | 29/02/20 | 07/03/20 |
| 2020-01-30 | 2.660 | 2.374 | null | >1.9 million | 01/09/20 | 11/03/20 |

<sup>a</sup>Date for the Wuhan city lockdown. <sup>b</sup>R0 before the lockdown. <sup>c</sup>R0 for the first month after the lockdown. <sup>d</sup>Simulated end point, day/month/year.
<sup>e</sup>Number of infected people at end point. <sup>f</sup>Simulated peak time, day/month/year. <sup>h</sup>Simulated turning time, day/month/year.

**Figure 3:**
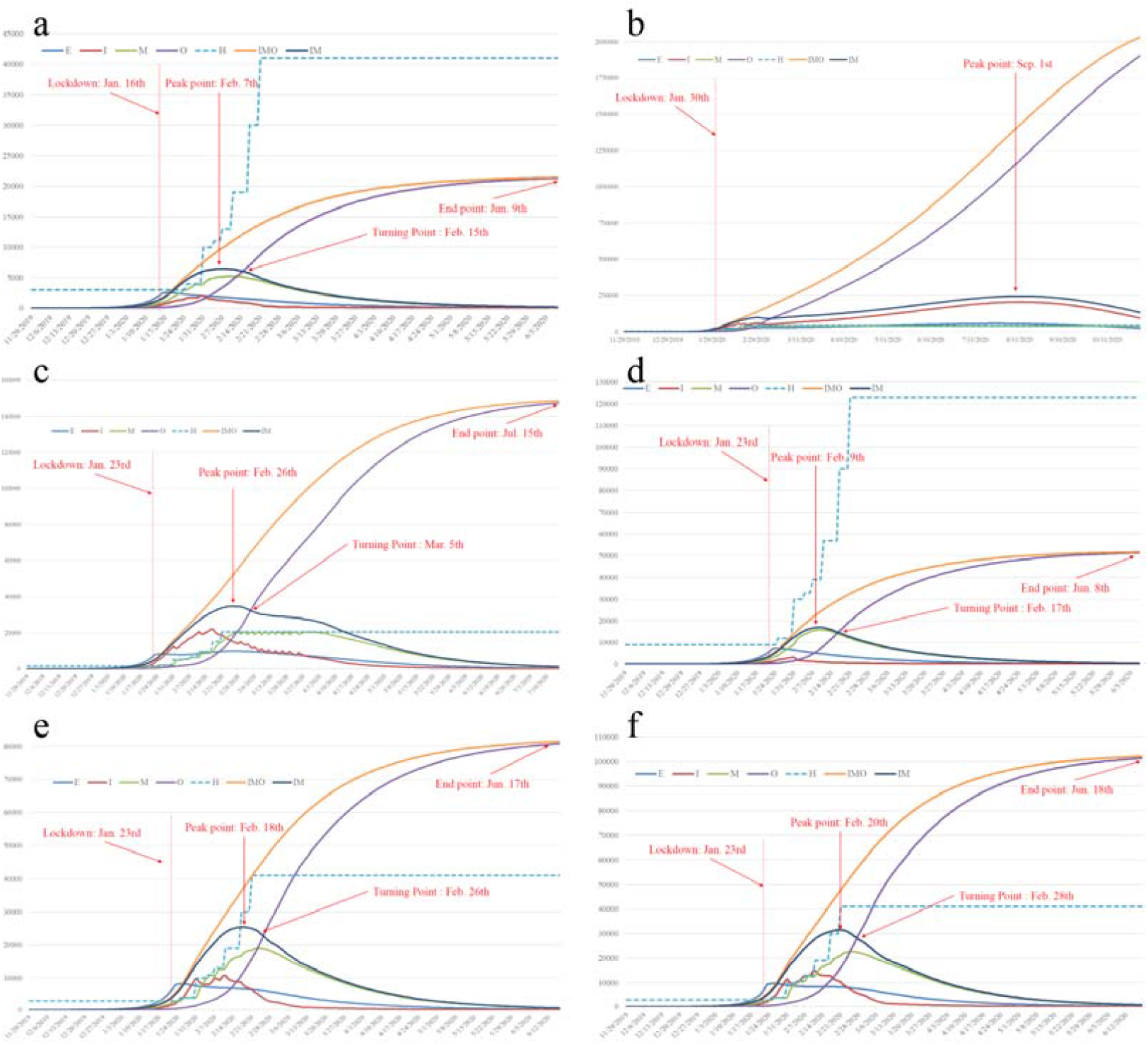
Status dynamic modeling of the epidemic under different public health response. a) Simulation results of locking down Wuhan 7 days earlier. b) Simulation results of 7 days delay for lockdown. c) Simulation results of 50% of beds in hospitals. d) Simulation of 300% of the current number of hospital beds. e) Simulation results of increasing 5% of parameter α on Jan 20^th^, 2020. f) Simulation results of increasing 25% of parameters α on Jan 20^th^, 2020.

On the contrary, if the lockdown was delayed by 1 to 7 days, the peak number would be dramatically increased. It could be found that, if the city was blocked 1 to 6 days later than Jan. 23^th^, 2020, the total infected people would be 1.23 times (95,273), 1.47 times (114,105), 1.83 times (141,409), 2.28 times (176,300), 2.90 times (222,384) and 4.94 times (382,695) than Jan. 23^th^, 2020 (Supplementary Figure 1 & Table 1), which was also consistent with one previous study ^14^. When the lockdown was 7 days later than the current choice, the first peak would arrive at Mar. 1^st^, 2020 with 97,779 infected people and the second peak would reach 243,693 on Sept. 1^st^, 2020. The number of total infected people would exceed 1.9 million and make the epidemic uncontrollable within the year 2020 (Figure 3b).

#### Impact of hospital bed numbers

Our simulation results showed that there was a shortage of hospital beds at the early stage of the epidemic, in which people in status *I* would not be completely isolated and treated with medical care. In that case, we simulated the effect of hospital bed numbers by multiplying its value by factors from 0.5 to 3 times, for each time point compared with current situations. Results indicated that if the beds in hospitals were halved, the peak time and endpoint would be Feb. 26^th^ and Jul. 15^th^, respectively. Moreover, 34,526 total infectious would be observed in the peak time, 1.43 times than the current situation, and 148,848 people would be infected eventually (Figure 3c). If the beds in hospitals were 1.5, 2, 2,5 and 3 times compared to the current situation, the total infectious number would be 62,353, 56,031, 53,245 and 51,898, respectively (Supplementary Table 1). Typically, tripling the beds in hospitals would end the epidemic 9 days earlier than the current situation (Figure 3d). Different from the lockdown of the city, when beds in hospitals were 2 times of current situations at each time point, the effect of increasing beds in hospitals was no longer significant (Supplementary Figure 2). In fact, when increasing the beds of hospitals by 3 times of the current situation, the total infectious at the endpoint would only decrease by 2.6% (1,347) than the 2.5-fold increase.

#### Impact of public gathering events

Since the time of the epidemic was overlapped with the Chinese Spring Festival and winter holiday, people were gathering together for celebration during the epidemic time. Thus, public gathering events would frequently happen if no prevention policies were imposed. For example, Wuhan conducted the banquet for ten thousand families on Jan 18^th^, 2020 ^20^, 5 days before the city lockdown. Here, we simulated the impact of public gathering events by increasing the transmission parameter α from 5% to 25% in an individual day such as Jan 20^th^, 2020.

Results showed that, if we increase α by 5% in one single day such as Jan 20^th^, the number of infections in peak time and end point will increase by 1,237 (5%) and 4,243(5%), respectively (Figure 3e). If α is increased by 25%, the number of people in peak time and endpoint will increase by 7,449 (31%) and 24,929 (32%), respectively (Figure 3f). Other results were also illustrated in Supplementary Figure 3 & Supplementary Table 2. Thus, the public gathering events in one single day could significantly impact the epidemic.

Generally, we believe the impact of locking down the city is more significant than the increasing number of beds in hospitals and public gathering events. The delay of city lockdown might easily make the epidemic out of control. Also, the public gathering events were inconducive for the prevention and control of the epidemic, since the slight increase of transmission parameter α would significantly affect the total infectious numbers at the end point. The number of beds in hospitals was essential in the early period of the epidemic when infected people increased rapidly, after the peak point, the beds would be enough for the increasing number of infected people.

### Simulation of no intervention and early time to release the Wuhan

Finally, we simulated the extreme situation that no intervention is performed. The circulation intensity in the same time period last year was derived from BAIDU migration data ^17^ to generated the α and β without any interventions. By setting the total number in human as 9 million ^21^ the results showed that the *R*_0_ would reach to 4.4 and over 7,697,484 people would be infected on Jun. 21^st^, 2019 (Figure 4a), which indicated that the epidemic was totally out of control without any intervention.

**Figure 4:**
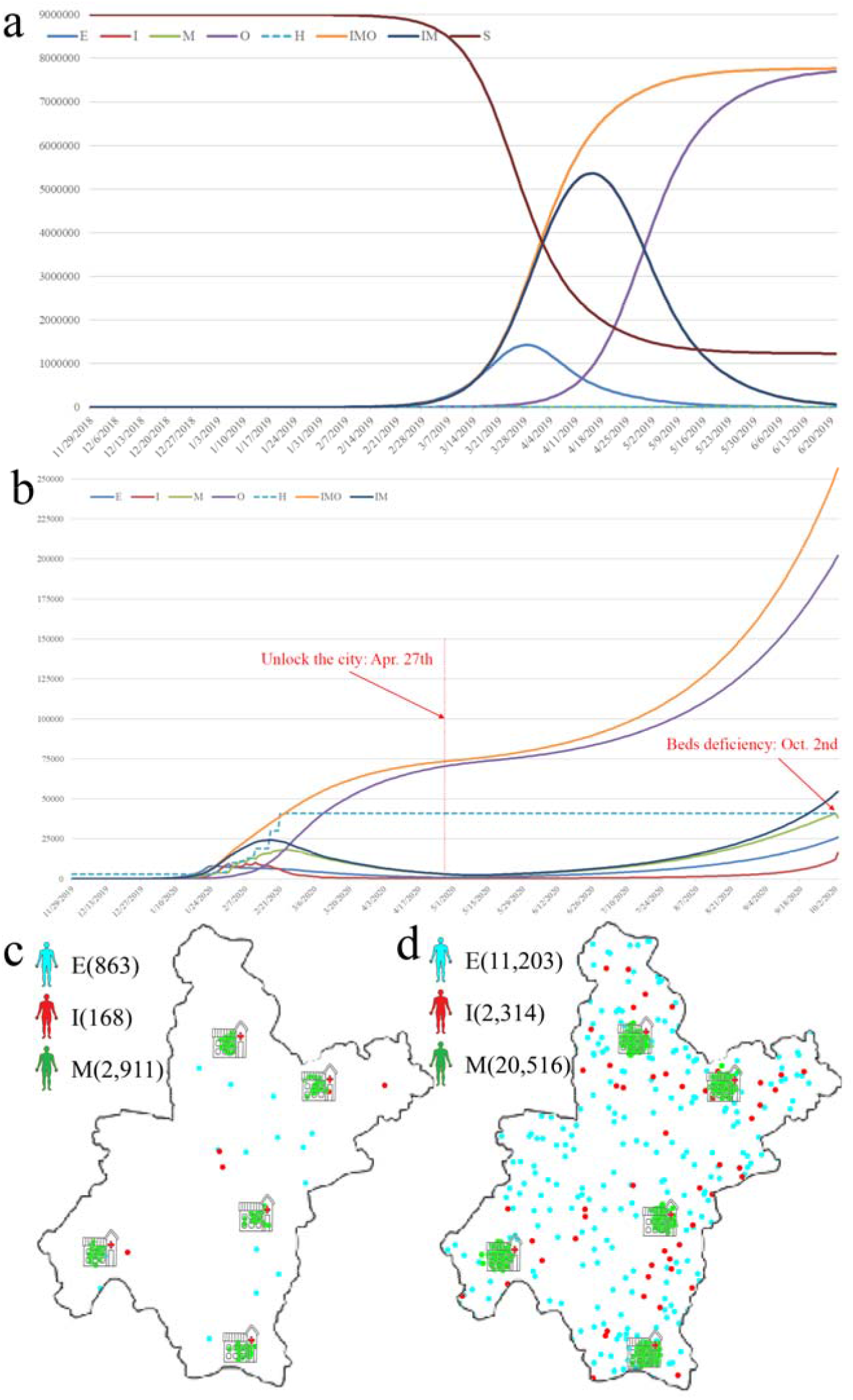
Status dynamic modeling of the epidemic without any intervention and early time to release Wuhan. a) Simulation results under no intervention in Wuhan based on the migration level in Wuhan last year. b) Simulation results for release Wuhan on Apr. 27^th^, 2020. c) The epidemic situation on Apr. 27^th^, 2020, 150 days after the lockdown. Blue, red and green dots located on the map of Wuhan represent the exposed, infected with symptoms and with medical care respectively. d) The epidemic situation on Jun. 26^th^, 2020, 60 days after the release of the city. Blue, red and green dots located on the map of Wuhan represent the exposed, infected with symptoms and with medical care respectively.

Another issue was about the early release of Wuhan. We experimented with the release time point of 150 days after the occurrence of patient zero, which is Apr. 27^th^, 2020 (Figure 4b). Results showed that, at that time the epidemic seemed to be under control with only 168 infected with symptoms and 2,912 with medical care on Apr. 27^th^, 2020 (Figure 4c) and kept decreasing until Mar 9^th^, 2020, in which the status number of infected with symptoms and 2,912 with medical care was 178 and 2,201 respectively. However, the number of exposed people increased from 863 on Apr. 27^th^ to 1,085 on Mar 9^th^, 2020, with the epidemic rapidly rebounded. After 120 days of releasing the city, on Aug. 25^th^, 2020, 11,203 exposed, 2,315 infected with symptoms and 20,516 with medical care were predicted (Figure 4d). The beds in hospitals would not be enough again on Oct. 2^nd^, 2020 with over 256,730 accumulated infections.

Although the transmission parameter α and β would not be the same as those before the lockdown, they were both increased by 25% (compared to the values during lockdown) to simulate the early release of the city. The results indicated that it was still a great risk to release the city before the epidemic reached the end point, even though the situation seemed to be under control.

## Discussion

The epidemic of SARS-CoV-2 has spread to China and multiple continents, which will be developed into a worldwide epidemic. During the epidemic period, nowcasting and forecasting are crucial for public health planning and control domestically and internationally ^12,22^. Here, we constructed a real-time status dynamic SEIO (MH) model to estimate the local outbreaks. By adding the parameters of status M and H, this model could accurately forecast the epidemics when the medical system under unexpected pressure. Moreover, this model could simulate the influence of different public health policies such as 1) lockdown of the city and 2) construction of temporary hospitals, which could help design the appropriate public health policies for epidemic control. At present, besides Wuhan, the epidemic situation in China has been effectively contained within a short period of time. This is largely due to the rapid response of the Chinese governments at various levels and the decision to lock down Wuhan, which is the first time to block a city with over 9 million permanent residents in human history. The lockdown of Wuhan would not only prevent the transportation between Wuhan to other cities, but also decrease the circulation intensity of people within Wuhan. According to BAIDU migration data, the circulation index of Wuhan is 6 to 8 times lower than those before the lockdown. The number of newly confirmed cases increased rapidly after the lockdown may due to the fact that, there have been already 4,521 infected and 6,536 exposed without symptoms before lockdown. It is noted that only 444 confirmed cases were reported by the Health Commission of Hubei Province ^16^. However, by facing an epidemic with the new pathogen, it is difficult to invent the diagnosis kit in such a short time. Thus, the diagnosis quantity might affect the early understanding of the epidemics and the lockdown decision at that time was decisive and timely. In fact, even one day delay of the lockdown would make 17,631 more infected people at the end point and a one-week delay would make the epidemic in Wuhan uncontrollable with over 1.9 million infectious.

Despite the advantage of epidemic control, the lockdown of Wuhan has significantly increased the pressure of local medical resources, including the rapidly growing demands for hospitalization, lack of medical staff such as doctors and nurses, and shortage of medical supplies. The simulation results illustrated the establishment of new temporary hospitals that could accommodate more patients could significantly help the control of the epidemic. In fact, if the beds in hospitals were three times more than the current situation, the total infectious would decrease by 33% at the end point. After the period of rapidly rising, beds in hospitals were not an essential parameter since the daily increase of infected people was less than the people out of the system. The lock of medical staff and the shortage of medical supplies increased the death rate in Wuhan, which was significantly higher than the rest of China. Based on this situation, multiple provinces sent medical teams and supplies to support Wuhan for epidemic control and treatment, which might help improve the curative ratio. Till Feb. 5^th^, 2020, nearly 7,000 medical staff were sent to Wuhan by governments at all levels in China ^23^.

Another noteworthy point is the public gathering events. Since the epidemic is overlapped with the Chinese Lunar New Year, public gathering events such as banquet of one thousand families in Wuhan could promote the spread of the epidemic. By adding a small turbulence (5%) in transmission parameter α on Jan 20^th^, 2020, 4,194 more people would be infected at the end point, which indicated the public gathering events at the early period of the epidemic should be stopped. Thus, the Chinese government prohibited the public gathering events with policies such as close large public places, extend the winter holiday for both staff and students. It is noted that, Japan and Koran also organized public gathering activities such as marathons, sacrifices activities, and rally in early February, 2020, which would be inconducive to the prevention and control of the epidemic.

The model forecasted that the peak time in Wuhan has been past and the epidemic in China is now under control. Currently, multiple countries including Koran, Japan, Iran, Italy, and the United States are in the outbreak stage ^24^. As the first country facing the SARS-CoV-2 epidemic, the experience of China in fighting the epidemics might provide useful guidance for other countries. According to our modeling study, we believe that the most effective public health policies for SARS-CoV-2 epidemic prevention are provided as follows: 1) reduce the local circulation intensity of people, 2) prepare enough medical resources, especially available beds in hospitals, 3) try to prohibit large public gathering events. It is expected that this study might provide useful guidance for other countries in fighting against the epidemic of COVID-19 or other similar infectious diseases.

## Data Availability

All code used in this study is available at https://github.com/xiaohan2012/covid-19-code.

## Contributors

TYQ collected the data and designed the model and experiment. HX formally defined the model and conducted the experiments. Both of them contributed to the writing of the manuscript.

